# The mass balance model perfectly fits both Hall *et al*. underfeeding data and Horton *et al*. overfeeding data

**DOI:** 10.1101/2021.02.22.21252026

**Authors:** Francisco Arencibia-Albite, Anssi H. Manninen

## Abstract

**Background & Aims:** Recently, the validity of mass balance model (MBM) was questioned based on two feeding studies. Thus, we simulated both of these feeding trials.

**Methods:** MBM describes the temporal evolution of body weight and body composition under a wide variety of feeding experiments. This computational study simulated, utilizing MBM, the underfeeding trial by Hall et al. (*Cell Metab*. 2015;22:427-36) and the overfeeding trial by Horton et al. (*Am J Clin Nutr*. 1995;62:19-29).

**Results:** Our simulation results indicate that data from both of these feeding trials perfectly match MBM-based predictions, i.e., MBM gives a remarkably accurate description of experimental data.

**Conclusions:** It is becoming increasingly clear that our model (MBM) is perfectly able to predict body weight and body composition fluctuations under a wide variety of feeding experiments.

“There is a stupid humility that is not at all rare, and those afflicted with it are altogether unfit to become devotees of knowledge. As soon as a person of this type perceives something striking, he turns on his heel, as it were, and says to himself: “You have made a mistake. What is the matter with your senses? This cannot, may not, be the truth.” And then, instead of looking and listening again, more carefully, he runs away from the striking thing, as if he had been intimidated, and tries to remove it from his mind as fast as he can. For his inner canon says: “I do not want to see anything that contradicts the prevalent opinion. Am I called to discover new truths? There are too many old ones, as it is…”

– Friedrich Nietzsche, *The Gay Science* (1882)

## Introduction

Currently, weight management is based on the “calories-in, calories-out” rule, formally named the energy balance theory (EBT). It maintains that body weight increases as food calories are greater than expended calories and *vice versa*. However, our recent papers [1,2] indicate that EBT-based simulations are *clearly erroneous*. As an alternative, the **mass balance model (MBM)** was proposed; and our recent simulations [1,2] indicate that MBM-based predictions are remarkably accurate.

Recently, the validity of mass balance model (MBM) was questioned based on two feeding studies, namely the underfeeding trial by Hall *et al*. [3] and the overfeeding study by Horton *et al*. [4]. Thus, we decided to simulate, utilizing MBM, both of the above mentioned feeding trials.

## Materials & Methods

### Mass balance model (MBM)

The MBM describes the temporal evolution of body weight and body composition under a wide variety of feeding experiments by using of the following measurements:

#### Energy-providing mass (EPM)

The daily mass intake given by carbohydrate, fat, protein, soluble fiber and alcohol.

#### Non-energy-providing mass (nEPM)

The daily mass intake given by insoluble fiber, vitamins, minerals and the net daily water balance (i.e., water ingestion and metabolically produced water minus lost water).

**R**: The relative daily rate of mass excretion free of total daily O_2_ uptake.

**β:** The mathematical formula that approximates the relationship between dFFM/dFM and FM is an *equilateral hyperbola* where β is the constant term of the hyperbola, i.e., FM (dFFM/dFM) = β. This parameter sets the magnitude of the change in fat free mass (FFM) whenever fat mass (FM) changes.

These measurements are used to populate the following MBM formulas:

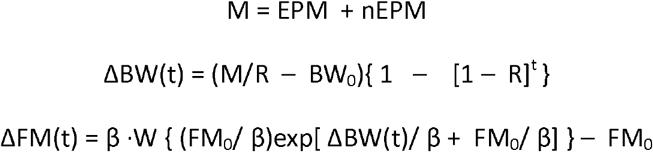

where t is the time in days; M is the total daily mass intake; BW_0_is the initial body weight; ΔBW(t) and ΔFM(t) are the cumulative body weight and fat mass changes, respectively; FM_0_ is the initial fat mass; and W is the product log function. For further details, consult the original MBM paper [1].

### MBM and Hall et al. underfeeding data

As was already pointed out by Arencibia-Albite in “Discussion” of the original MBM paper [1], this model, in fact, perfectly fits Hall *et al*. data [3]. In their Table 3 and Figure 3F, it is shown that a period of six days of reduced carbohydrate diet (RC) resulted, on average, in a total weight loss of –1.85kg, whereas the reduced fat diet (RF) resulted in –1.3kg (RC vs. RF, p=0.022). According to their Table 1, EPM of each diet is as follows:

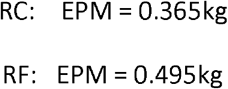

while the average BW_0_ and FM_0_ are 106kg and 42kg, respectively.

Using this data and standard numerical methods, we approximated that the remaining MBM parameters were:

| Parameters | RC | RF |
| --- | --- | --- |
| **nEPM** | 1.4846kg | 1.6055kg |
| **R** | 0.0205109 | 0.0219753 |
| **B** | 104.2324kg | 50.5016kg |

Therefore, the MBM formulas for each diet are as follows:

RC:

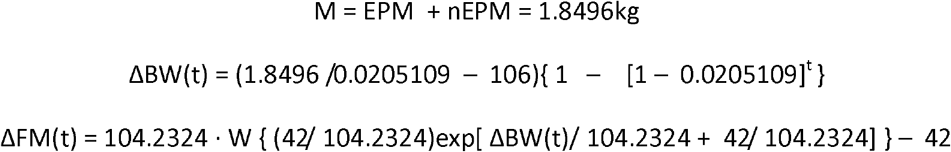

RF:

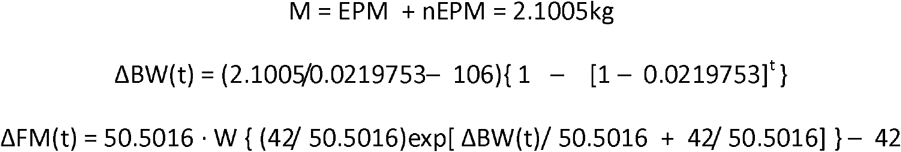

These formulas are plotted in **Figure 1**. For further details, see the figure legend.

**Figure 1.**
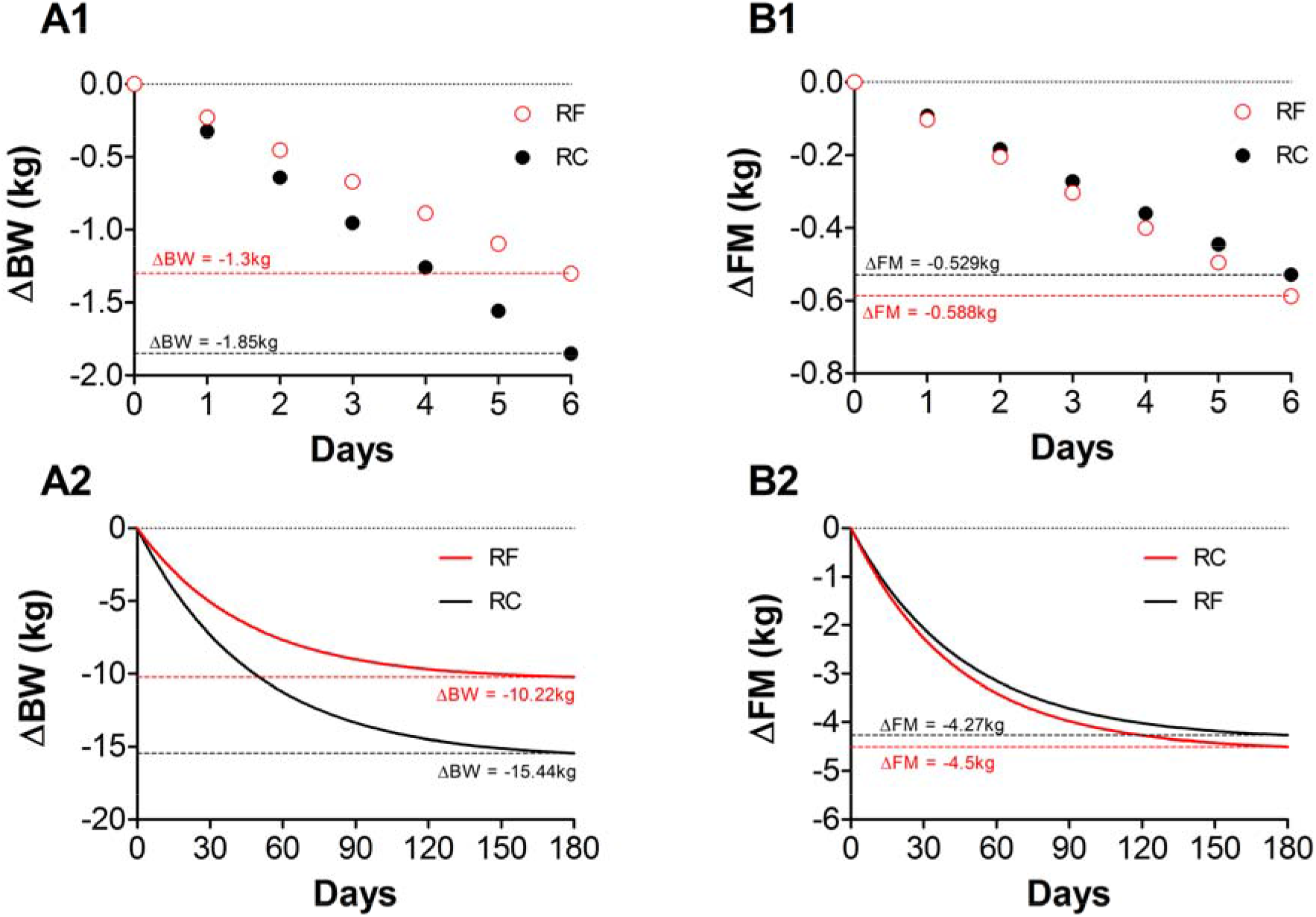
MBM simulation of Hall et al. underfeeding data. **A1**. MBM weight loss trajectories perfectly match those reported by Hall *et al*. [3] over the trial duration (6 days). **A2**. The graph shows the same weight loss trajectories as in panel A1 but extended for 180 days. The MBM predicts a greater weight loss in the RC diet in contrast to the RF diet. **B1**. Fat loss trajectories that underlay weight loss in A1 perfectly match those reported by Hall *et al*. [3] over the trial duration (6 days). **B2**. The graph shows the same fat loss trajectories as in panel B1 but extended for 180 days. The MBM predicts very similar levels of fat loss between diets.

### MBM and Horton *et al.* overfeeding data

According to Table 1 in Horton *et al*. [4], the average initial characteristics of the lean and obese subjects were as follows:

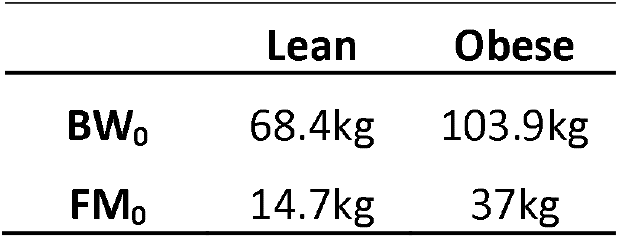

After an isocaloric overfeeding period of 14 days, the latter characteristics change (Table 2 in Horton *et al*. [4]):

|  | Lean |  | Obese |  |
| --- | --- | --- | --- | --- |
|  | High Carb. (HC) | High Fat (HF) | High Carb. (HC) | High Fat (HF) |
| **ΔBW** | 2.47kg | 2.31kg | 3.47kg | 2.98kg |
| **ΔFM** | 1.09kg | 1.21kg | 2.06kg | 1.90kg |

The associated EMPs that lead to these changes were as follows:

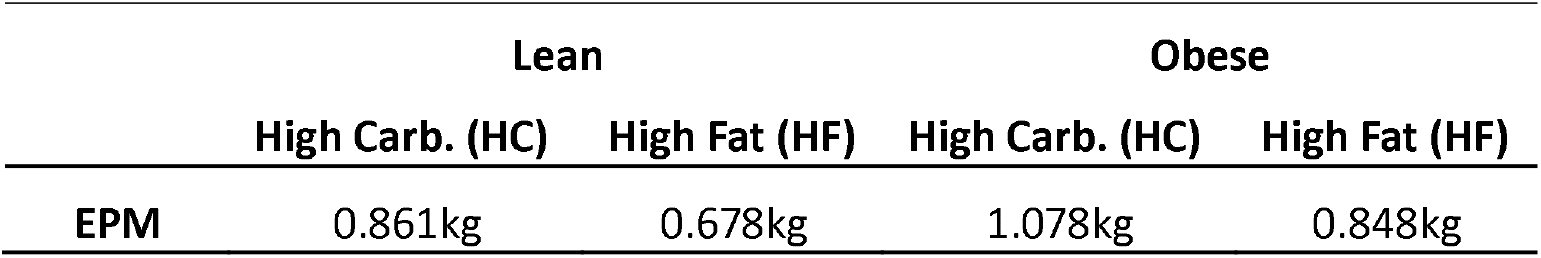

|  | Lean |  | Obese |  |
| --- | --- | --- | --- | --- |
|  | High Carb. (HC) | High Fat (HF) | High Carb. (HC) | High Fat (HF) |
| **EPM** | 0.861kg | 0.678kg | 1.078kg | 0.848kg |

Using the above data and standard numerical methods, we approximated that the remaining MBM parameters were:

|  | Lean |  | Obese |  |
| --- | --- | --- | --- | --- |
|  | High Carb. (HC) | High Fat (HF) | High Carb. (HC) | High Fat (HF) |
| **nEPM** | 0.650kg | 0.820kg | 0.800kg | 1.020kg |
| **R** | 0.019174 | 0.019173 | 0.01544 | 0.0157126 |
| **β** | 19.29597kg | 13.90792kg | 26.04379kg | 21.56814kg |

Therefore, the MBM formulas for each diet are as follows:

Lean HC:

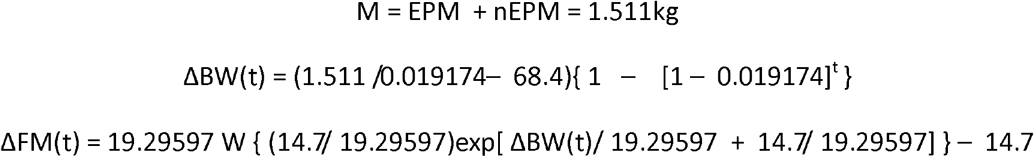

Lean HF:

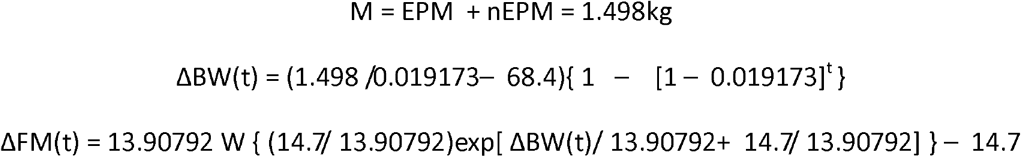

Obese HC:

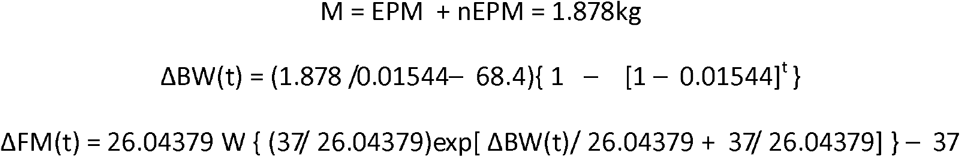

Obese HF:

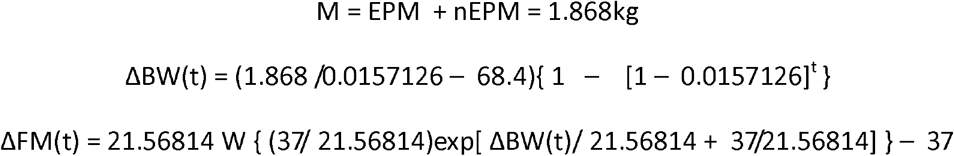

All the above formulas are plotted in **Figure 2**. For further details, see the figure legend.

**Figure 2.**
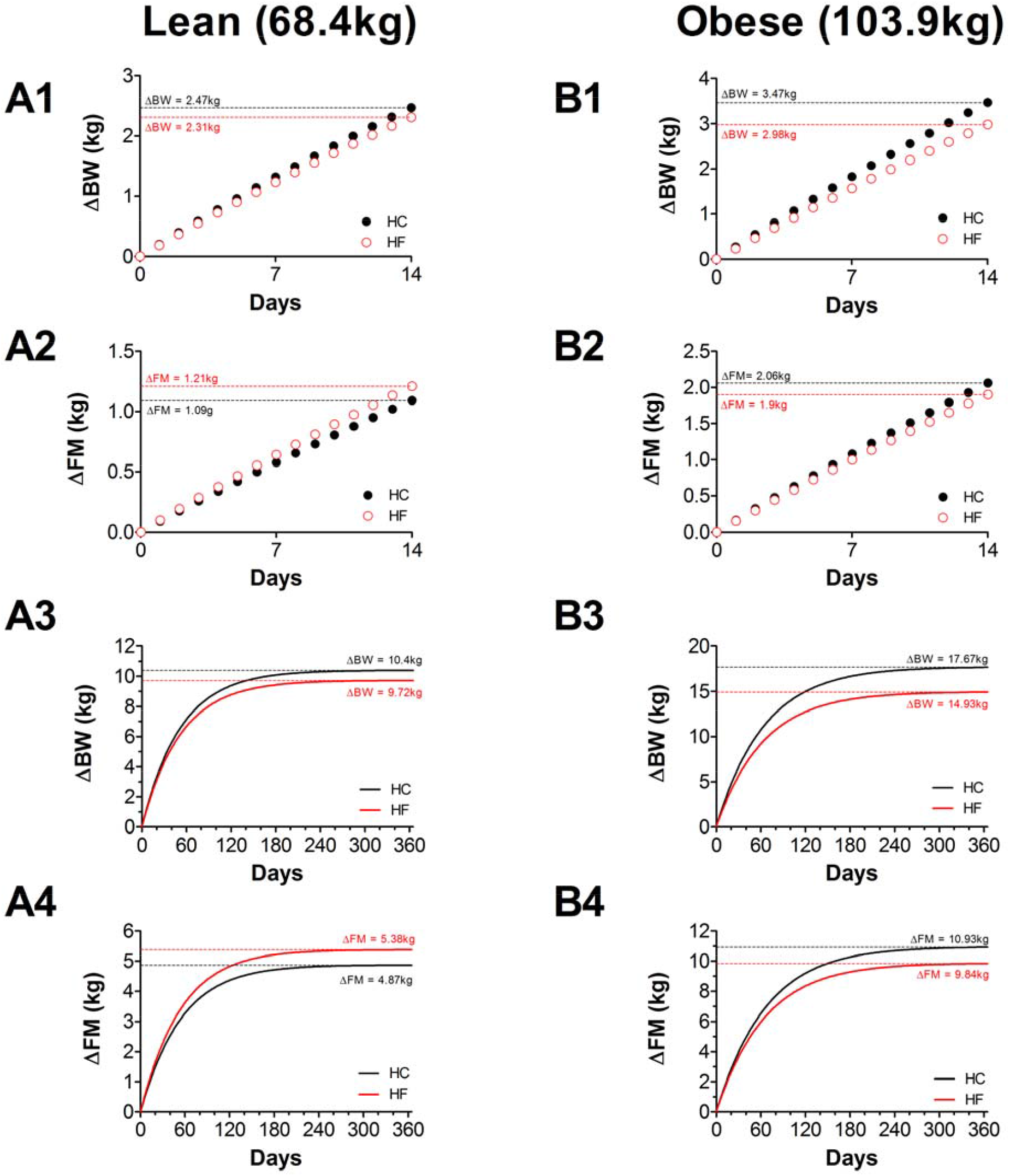
MBM simulation of Horton et al. overfeeding data. **A1**. MBM weight gain trajectories for the lean subjects perfectly match those reported by Horton *et al*. [4] over the trial duration (14 days). **A2**. Fat gain trajectories that underlay weight gain in A1 perfectly match those reported by Horton *et al*. [4] over the trial duration (14 days). Notice that the MBM predicts very similar levels of fat gain among diets over the 14 days. **A3**. The graph shows the same weight gain trajectories as in panel A1 but extended for 365 days. **A4**. The graph shows the same fat gain trajectories as in panel A2 but extended for 365 days. **B1**. MBM weight gain trajectories for the obese subjects perfectly match those reported by Horton *et al*. [2] over the trial duration (14 days). **B2**. Fat gain trajectories that underlay weight gain in B1 perfectly match those reported by Horton *et al*. [2] over the trail duration (14 days). Notice that the MBM predicts very similar levels of fat gain among diets over the 14 days. **B3**. The graph shows the same weight gain trajectories as in panel B1 but extended for 365 days. **B4**. The graph shows the same fat gain trajectories as in panel B2 but extended for 365 days. HC = high-carbohydrate diet; HF = high-fat diet.

## Discussion

Recently, the validity of MBM was questioned [5] based on two feeding studies [3,4]. Thus, we decided to simulate both of these feeding trials [3,4]. Our “mass in, mass out” model (MBM) describes the temporal evolution of body weight and body composition under a wide variety of feeding experiments.

If *1)* nEPM, *2)* R, and *3)* β are the same between the diets, the diet with the greatest macronutrient mass will lead to greatest weight and fat gain during overfeeding, whereas the diet with the smallest macronutrient mass will result in greatest weight and fat loss during underfeeding. When these parameters (i.e., nEPM, R, β) are taking into consideration, MBM gives a remarkably accurate description of experimental data under a wide variety of feeding experiments (See also [1,2]).

## Conclusion

Our results indicate that MBM is perfectly able to predict body weight and body composition fluctuations under a wide variety of feeding experiments. Thus, researchers around the world should acknowledge that the widely accepted <u>EBT is clearly erroneous</u> (See also [1,2]).

## Data Availability

N/A

## Authors contribution

Study concept and designing: Manninen, Arencibia-Albite

Acquisition of data: Manninen, Arencibia-Albite

Statistical analysis and interpretation of data: Manninen, Arencibia-Albite

Drafting the manuscript: Manninen, Arencibia-Albite

Critical revision of the manuscript for important intellectual content: Manninen, Arencibia-Albite

Administrative, technical or material support: Manninen, Arencibia-Albite.

Study supervision: N/A.

## Declaration of competing interests

The authors declare that they have no competing interests.

### List of abbreviations

MBM: mass balance model
EBT: energy balance theory
BW: body weight
FM: fat mass
FFM: fat free mass
EPM: energy-providing mass
nEPM: non-energy-providing mass
RC: carbohydrate restricted diet
RF: fat restricted diet
HC: high-carbohydrate diet
HF: high-fat diet.

